# Accelerometer-measured habitual physical activity and sedentary time in pediatric concussion: A controlled cohort study

**DOI:** 10.1101/2021.07.15.21260582

**Authors:** Bhanu Sharma, Joyce Obeid, Carol DeMatteo, Michael D. Noseworthy, Brian W. Timmons

## Abstract

**Objectives:** To characterize and quantify differences in accelerometer-measured physical activity and sedentary time between children with concussion (within the first month of injury) and 1:1 matched healthy controls.

**Methods:** Secondary analysis of accelerometer data collected on 60 children with concussion and 60 healthy controls matched for age, sex, and season of accelerometer wear. Daily and hourly sedentary time, light physical activity (LPA), moderate physical activity (MPA), and vigorous physical activity (VPA) were compared between groups per independent samples t-tests.

**Results:** Children with concussion (12.74 ± 2.85 years, 31 females) were significantly more sedentary than controls (12.43 ± 2.71 years, 31 females; mean difference [MD], 38.3 minutes/day, p=0.006), and spent less time performing LPA (MD, -19.5 minutes/day, p=0.008), MPA (MD, -9.8 minutes/day, p<0.001), and VPA(MD, -12.0 minutes/day, p<0.001); hour-by-hour analyses showed that these differences were observed from 8:00AM to 9:00PM. Sex-specific analyses identified that girls with concussion were less active and more sedentary than both boys with concussion (MD, 50.8 minutes/day; p=0.010) and healthy girls (MD, 51.1 minutes/day; p<0.010). Days post-injury significantly predicted MPA (β=0.071, p=0.032) and VPA (β=0.095, p=0.004), but not LPA or sedentariness in children with concussion.

**Conclusion:** Clinical management should continue to advise against prolonged rest following pediatric concussion, given the activity debt observed within the first-month of injury. Currently, clinical management of concussion is shifting towards prescribing a single bout of daily sub-maximal aerobic exercise. Interventions aimed at reducing overall sedentary time and increasing habitual physical activity in pediatric concussion also warrant study.

**KEY FINDINGS:** *What are the new findings?:*

- Per accelerometry, children with concussion are significantly more sedentary than healthy controls within the first-month of injury
- Relative to healthy controls, in the first-month of injury, children with concussion perform less accelerometer-measured light, moderate, and vigorous physical activity
- These patterns of increased sedentary time and reduced physical activity are seen throughout the day, from 8:00 AM to 9:00 PM
- Moderate and vigorous physical activity levels are predicted by days-post injury in children with concussion, and may increase naturally as a consequence of recovery

*How might it impact on clinical practice in the future?:*

- Given the accumulating evidence that prolonged rest should be avoided following concussion, primary care clinicians should continue to advocate for light physical activity post-concussion to limit increased sedentary time

## INTRODUCTION

Concussions are brain injuries that are caused by biomechanical impact and result in functional neurological disturbance^1^. These injuries are particularly common in pediatric populations where the incidence of concussion is rising^2-5^. While the majority of pediatric concussion cases resolve within an expected four-week window^1^, nearly 30% may result in long-term symptoms^6-8^. Delayed recovery can lead to poor academic outcomes^9^, health-related quality of life^10-12^, and mental health^13^.

Accordingly, research into an effective treatment for concussion has become a priority. Sub-maximal aerobic exercise has recently emerged as a leading candidate for the treatment of concussion symptoms^14-16^. The rapidly accumulating evidence on the benefits of exercise in concussion has been summarized by multiple meta-analyses^17-19^, the most recent of which examined 23 studies (N=2547), finding that sub-maximal aerobic exercise has a large, positive effect (*Hedges’ g*=1.71) on concussion recovery^17^. A paradigm shift in concussion management is occurring, wherein sub-maximal aerobic exercise is supplanting prolonged rest as a management strategy in both pediatric and adult concussion^15,20,21^.

However, levels of habitual physical activity and sedentary time in children with concussion have not been studied, and we do not yet know whether these levels differ from their typically developing peers or whether they vary by sex (as do many other clinical features of concussion^22,23^). In a field where the deleterious impacts of prolonged rest on recovery are being increasingly recognized^19,24,25^, a lack of knowledge of baseline physical activity and sedentary time after pediatric concussion represents a critical knowledge gap. Not only are we unaware of whether there is a potentially symptom prolonging physical activity deficit that needs to be overcome in pediatric concussion, but we also do not know of the impact of concussion on a critical aspect of daily functioning, namely participation in routine activity. In a cohort of adults with concussion (n=180), per a self-recall questionnaire, 85% identified as meeting physical activity guidelines pre-injury, compared to 28% post-concussion^26^. Similar data are not available on children. Such data, however, create opportunity to examine whether current exercise interventions aimed at providing a once-a-day aerobic stimulus are adequate^14-16^, or if interventions that increase levels of habitual physical activity and reduce overall sedentariness throughout the day are warranted.

Habitual physical activity can be objectively quantified using accelerometers^27,28^, which permit high temporal resolution quantification of movement by measuring multi-axis accelerations. Accelerometers have been used in many pediatric neurological and chronic disease populations to compare the activity of these children to their typically developing peers^29-31^, providing insights about the intensity, time, and frequency of physical activity that recall surveys cannot capture. In concussion, accelerometry has been used to begin to understand the relationship between concussion symptoms and activity^32-34^, although no controlled studies have quantified levels of sedentary time and habitual physical activity.

This study was performed to characterize accelerometer-measured physical activity patterns of children with concussion in comparison to their healthy peers. Learning more about habitual physical activity and sedentary time within the first month after concussion can aid in clinical management and informing the next wave of exercise intervention research.

## METHODS

This study was approved by the Hamilton Integrated Research Ethics Board (www.hireb.ca).

### Design

This study is a secondary analysis of accelerometer data collected as part of a prospective cohort study (led by the senior authors) aimed at developing protocols for safe resumption of activity following pediatric concussion.

### Participants

Children (aged 6-17) presenting acutely to the emergency department at McMaster Children’s Hospital, or to affiliated community physicians and sports medicine clinics, who received a diagnosis of concussion were referred to the research team. Informed consent from parents and assent/consent (as appropriate) from teenagers was obtained. Exclusion criteria were more severe brain injury or complex injuries involving multiple organ systems, and clinically diagnosed neurological or developmental disorder.

Participants with a concussion were matched 1:1 to healthy controls. The healthy controls were children who participated in prior studies in our lab, either as participants in longitudinal physical activity studies of healthy children or as controls of children from clinical populations. Patients with concussion were matched to healthy controls on three criteria, namely chronological age, sex, and season/month the accelerometer was worn. Matching for season/month (within 60 days) was important given known accelerometer-measured seasonal variation in both sedentary and active time in children^35^.

### Procedures

At initial intake, participants were provided with a waist-worn accelerometer to measure habitual physical activity. More specifically, the accelerometer used was the ActiGraph GT3x (Pensacola, FL, USA), a tri-axial axes accelerometer that is small, light-weight and unobtrusive during daily wear. This unit has also been shown to measure physical activity in acquired brain injury with high reproducibility^36^. The accelerometer was set to record movement at 30 Hertz. Participants were instructed to wear the accelerometer on their right hip during waking hours, except when engaged in water-based activities. Participants with concussion were asked to wear the device throughout their recovery, up to a maximum of 6 months; the current study focuses on the first 4 weeks, or the expected timeframe for recovery. Healthy controls were asked to wear the device for 7 to 9 consecutive days. Further, participants were given a logbook and instructed to note every time the device was taken off and re-worn.

All accelerometer data were downloaded in 3-second epochs and processed in ActiLife Software (ActiGraph; Pensacola, FL, USA). A semi-automated data cleaning procedure was used to detect any periods ≥5 minutes of zeros. Each of these bouts were inspected and only non-wear periods identified using the participant logbooks were excluded from subsequent data processing. Days with missing logbook entries were also excluded from the analysis. The manually cleaned data were then scored to determine activity by intensity, using the validated Evenson cut points^37^. These cut points were scaled to 3-sec epoch data, and included time spent in sedentary time (0-25 counts/15s), light physical activity (LPA; 26-573 counts/15s), moderate physical activity (MPA; 574-1002 counts/15s), and vigorous physical activity (VPA; 1003+ counts/15s).

A sedentary bout analysis was also performed to quantify the length of individual sedentary bouts and the time between them. Sedentary bouts were defined as at least 1 epoch (3-sec) in length with an activity count ≤100 counts per minute, with a drop time of no more than 2-epochs. This meant that within any given sedentary bout, up to 6-sec of non-sedentary time was ignored (i.e., dropped), which would be akin to a positioning adjustment or reaching for a remote control or phone.

### Data analysis

Data were imported into SPSS Version 27 (IBM Corp. Released 2020. IBM SPSS Statistics for Windows, Version 27.0. Armonk, NY). Minimum wear time criteria were ≥ 600 minutes of for ≥ 4 days^38^. For hour-by-hour analyses, we included only hours with ≥ 50 minutes of wear time to ensure that the data were representative of the activities performed in the hour. Once the final sample of participants with valid wear data was acquired, daily average and hour-by-hour average activity and sedentary time was computed at the single-subject and group levels. Normality was assessed using Shapiro-Wilk tests, and independent sample t-tests were then used to compare accelerometer data by group, at both the daily and hourly levels. More specifically, sedentary time, LPA, MPA, and VPA were compared between groups, with Bonferroni corrections to adjust for family-wise multiple comparisons. Further, we examined the association between sedentary time and activity levels with days post-injury. General linear models (GLMs) were developed (to understand activity as a function of time post-injury) with sedentariness and activity levels as outcomes, and days post-injury and age as predictors; distribution of residuals was subsequently tested.

## RESULTS

### Overview

Our sample was comprised of 60 children with concussion and 60 healthy controls. Children with concussion wore their accelerometers for significantly longer overall; however, wear time per day was not significantly different between groups. These data along with demographics and the number of days and hours dropped from the analysis for not meeting wear time requirements, per group, are reported in **Table 1**.

**Table 1.**
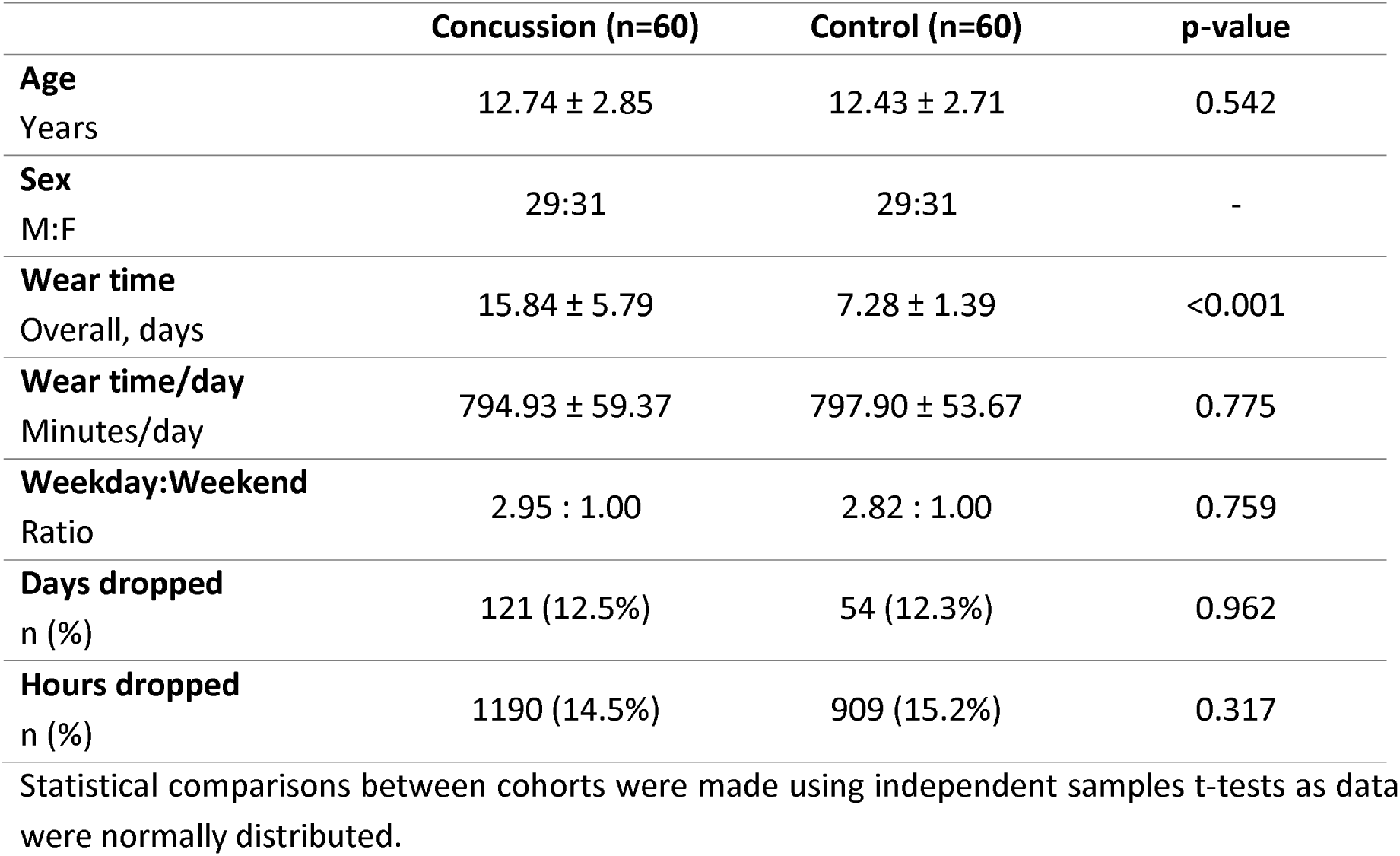
Demographic and wear time data for the study.

|  | **Concussion (n=60)** | **Control (n=60)** | **p-value** |
| --- | --- | --- | --- |
| **Age**<br>Years | 12.74 ± 2.85 | 12.43 ± 2.71 | 0.542 |
| **Sex**<br>M:F | 29:31 | 29:31 | - |
| **Wear time**<br>Overall, days | 15.84 ± 5.79 | 7.28 ± 1.39 | <0.001 |
| **Wear time/day**<br>Minutes/day | 794.93 ± 59.37 | 797.90 ± 53.67 | 0.775 |
| **Weekday:Weekend**<br>Ratio | 2.95 : 1.00 | 2.82 : 1.00 | 0.759 |
| **Days dropped**<br>n (%) | 121 (12.5%) | 54 (12.3%) | 0.962 |
| **Hours dropped**<br>n (%) | 1190 (14.5%) | 909 (15.2%) | 0.317 |

Statistical comparisons between cohorts were made using independent samples t-tests as data were normally distributed.

### Groupwise activity and inactivity differences

Sedentary time was higher in children with concussion in comparison to healthy controls (mean difference [MD], 38.3 minutes/day; p=0.006). Accordingly, LPA (MD, -19.5 minutes/day; p=0.008), MPA (MD, -9.8 minutes/day; p<0.001), and VPA (MA, -12.0 minutes/day; p<0.001) were significantly lower in children with concussion relative to controls (**Table 2**). Girls with concussion were significantly more sedentary than boys with concussion (MD, 50.8 minutes/day; p=0.010) and less active with respect to LPA (MD, -25.0 minutes/day; p=0.010) and MPA (MD, -8.7 minutes/day; p=0.004). There were no differences between healthy boys and girls. However, girls with concussion were significantly more sedentary and less active, across all intensities, than healthy girls (**Table 3**).

**Table 2.** Group-wise sedentariness and activity levels.

|  |  | **Concussion,**<br>**mean ± SD**<br>**(n=60)** | **Control,**<br>**mean ± SD**<br>**(n=60)** | **Mean**<br>**difference,**<br>**95% CI** | **t-stat**<br>**(118 df)** | **p-value** |
| --- | --- | --- | --- | --- | --- | --- |
| **Minutes/day** | Sedentary | 618.8<br>± 77.2 | 580.5<br>± 72.7 | 38.3<br>(11.2, 65.4) | 2.800 | 0.006* |
| | LPA | 130.2<br>± 42.9 | 149.7<br>± 35.2 | -19.5<br>(-5.3, -33.7) | -2.717 | 0.008* |
| | MPA | 27.2<br>± 10.6 | 37.0<br>± 11.7 | -9.8<br>(-5.7, -13.8) | -4.799 | <0.001* |
| | VPA | 18.7<br>± 12.6 | 30.7<br>± 15.7 | -12.0<br>(-6.9, -17.2) | -4.362 | <0.001* |

\*denotes significant findings at Bonferroni-corrected p-value of $\alpha/4$ (0.0125).

**Table 3.** Sex-based differences in sedentariness and activity between groups.

|  |  | Children with Concussion |  |  | Healthy Controls |  |  |
| --- | --- | --- | --- | --- | --- | --- | --- |
|  |  | Boys<br>(n=29) | Girls<br>(n=31) | t-stat<br>p-value | Boys<br>(n=29) | Girls<br>(n=31) | t-stat<br>p-value |
| **Minutes/day** | **Sedentary<sup>a</sup>** | 592.5 | 643.3 | -2.677 | 567.0 | 592.2 | -1.347 |
|  |  | ± 60.2 | ± 83.9 | 0.010* | ± 68.2 | ± 75.5 | 0.183 |
|  | **LPA<sup>a</sup>** | 143.1 | 118.1 | 2.335 | 150.8 | 148.7 | 0.226 |
|  |  | ± 36.7 | ± 45.3 | 0.012* | ± 25.8 | ± 42.2 | 0.822 |
|  | **MPA<sup>a</sup>** | 32.2 | 23.5 | 3.024 | 38.5 | 35.7 | 0.920 |
|  |  | ± 9.7 | ± 10.1 | 0.004* | ± 12.0 | ± 11.5 | 0.361 |
|  | **VPA<sup>a</sup>** | 22.3 | 15.3 | 2.367 | 34.3 | 27.6 | 1.672 |
|  |  | ± 10.6 | ± 13.5 | 0.011* | ± 16.3 | ± 14.7 | 0.100 |

\*denotes significant findings at Bonferroni-corrected p-value of $\alpha/4$ (0.0125). <sup>a</sup> Significantly different from healthy girls (p<0.0125).

Hourly analyses showed that from 8:00 AM to 9:00 PM, children with concussion were consistently more sedentary and less active than their healthy peers (**Figure 1**). Data from the earlier hours (i.e., before 8:00 AM) were not including in the analysis owing to limited availability of valid accelerometer data during this time. The effect sizes for the differences in hour-by-hour accelerometer-measured sedentary time and activity between children with concussion and healthy controls are presented in **Figure 1**.

**Figure 1.**
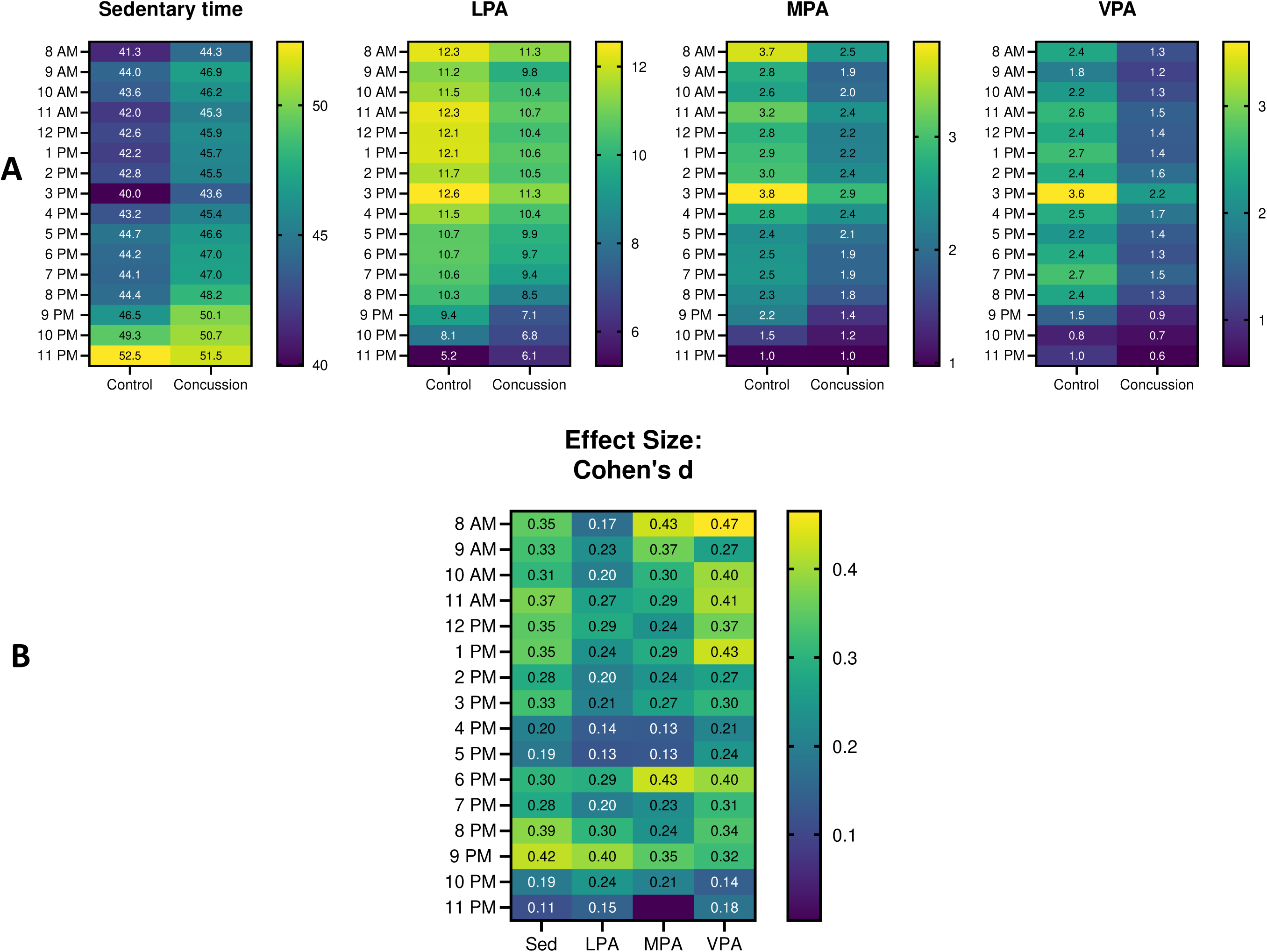
*<u>Panel A</u>*. Hour-by-hour sedentariness and activity (from 8:00 AM to 12:00 AM) in children with concussion in comparison to healthy controls. The numbers within the cells represent the average number of minutes of sedentariness or activity per hour (all differences significant except the hours of 10PM and 11PM). The bars represent minutes/hour. *<u>Panel B</u>*. The effect sizes (*Cohen’s d*) associated with the comparisons presented in Figure 1. Lighter cell colours represent larger effect sizes. (LPA = light physical activity; MPA = moderate physical activity; VPA = vigorous physical activity; ns = non-significant).

### Sedentary bout analysis

Our sedentary bout analysis revealed that healthy children had significantly more short sedentary bouts/hour (<1 minute) than children with concussion. However, children with concussion engaged in a significantly greater number of 5-to-10-minute sedentary bouts/hour than healthy controls (**Tables 4** and **5**).

**Table 4.** Group-wise sedentary bout analysis.

|  |  | **Concussion,**<br>**mean ± SD**<br>**(n=60)** | **Control,**<br>**mean ± SD**<br>**(n=60)** | **Mean**<br>**difference,**<br>**95% CI** | **t-stat**<br>**(118 df)** | **p-value** |
| --- | --- | --- | --- | --- | --- | --- |
| **Sedentary**<br>**bouts/hour** | <1<br>minute | 31.02<br>± 11.9 | 37.17<br>± 12.5 | -6.15<br>(-1.7, -10.6) | -2.762 | 0.003* |
| | 1- to 5-<br>minutes | 9.65<br>± 1.8 | 9.76<br>± 1.6 | -0.11<br>(-0.50, 0.71) | -0.359 | 0.720 |
| | 5- to 10-<br>minutes | 1.24<br>± 0.3 | 1.12<br>± 0.3 | 0.13<br>(0.01, 0.24) | 2.492 | 0.007* |
| | 10- to 20-<br>minutes | 0.45<br>± 0.2 | 0.38<br>± 0.2 | 0.07<br>(-0.01, 0.15) | 1.910 | 0.059 |
| | 20- to 30-<br>minutes | 0.08<br>± 0.06 | 0.06<br>± 0.06 | 0.02<br>(-0.01, 0.03) | 1.326 | 0.187 |
| | 30- to 60-<br>minutes | 0.04<br>± 0.04 | 0.03<br>± 0.04 | 0.01<br>(-0.01, 0.03) | 1.598 | 0.113 |
| **Breaks**<br>**between bouts** | <1<br>minute | 40.8<br>± 12.2 | 46.2<br>± 12.2 | -5.4<br>(-0.9, -9.8) | 2.420 | 0.017 |
| | 1- to 5-<br>minutes | 1.6<br>± 0.7 | 2.1<br>± 0.9 | -0.6<br>(-0.3, -0.9) | 3.959 | 0.000* |
| | 5- to 10-<br>minutes | 0.05<br>± 0.05 | 0.09<br>± 0.07 | -0.04<br>(-0.02, -0.06) | 3.476 | 0.001* |
| | 10- to 20-<br>minutes | 0.01<br>± 0.01 | 0.03<br>± 0.03 | -0.02<br>(-0.00, -0.02) | 3.246 | 0.002* |
| | 20- to 30-<br>minutes | 0.01<br>± 0.01 | 0.01<br>± 0.01 | 0.00<br>(0.00, 0.01) | 0.07 | 0.946 |
| | 30- to 60-<br>minutes | 0.01<br>± 0.01 | 0.01<br>± 0.01 | 0.00<br>(0.00, 0.01) | 1.357 | 0.177 |

\*denotes significant findings at Bonferroni-corrected p-value of $\alpha/6$ (0.008).

**Table 5.** Sedentary bout analysis, by sex and group.

|  |  | Children with Concussion |  |  | Healthy Controls |  |  |
| --- | --- | --- | --- | --- | --- | --- | --- |
|  |  | Boys<br>(n=29) | Girls<br>(n=31) | t-stat<br>p-value | Boys<br>(n=29) | Girls<br>(n=31) | t-stat<br>p-value |
| **Sedentary**<br>**bouts/hour** | <1 minute<sup>a</sup> | 33.84<br>± 10.0 | 28.38<br>± 13.0 | 1.815<br>0.075 | 35.42<br>± 11.3 | 38.69<br>± 13.5 | -1.012<br>0.316 |
|  | 1- to 5-minutes | 10.37<br>± 1.5 | 8.98<br>± 1.9 | 3.210<br>0.002* | 9.66<br>± 1.9 | 9.85<br>± 1.2 | -0.483<br>0.631 |
|  | 5- to 10-minutes | 1.23<br>± 0.3 | 1.26<br>± 0.3 | -0.384<br>0.702 | 1.14<br>± 0.3 | 1.10<br>0.4 | 0.550<br>0.585 |
|  | 10- to 20-<br>minutes | 0.41<br>± 0.2 | 0.49<br>± 0.2 | -1.460<br>0.150 | 0.39<br>± 0.2 | 0.37<br>± 0.2 | 0.479<br>0.634 |
|  | 20- to 30-<br>minutes | 0.06<br>± 0.05 | 0.09<br>± 0.06 | -2.240<br>0.029 | 0.07<br>± 0.06 | 0.06<br>± 0.05 | 0.708<br>0.489 |
|  | 30- to 60-<br>minutes | 0.03<br>± 0.04 | 0.05<br>± 0.04 | -2.023<br>0.048 | 0.03<br>± 0.01 | 0.02<br>± 0.01 | 0.327<br>0.745 |

\*denotes significant findings at Bonferroni-corrected p-value of $\alpha/6$ (0.008). <sup>a</sup> Significantly different ( $p < 0.008$ ) from healthy girls.

### Activity and inactivity as a function of days post-injury

GLMs predicting activity levels as a function of days post-injury and age were only significant for models (unstandardized betas reported) with MPA (β_Days Post_=0.177, t_2,842_=2.432, p=0.015) and VPA (β_Days Post_=0.265, t_2,842_=3.227, p=0.001) as the outcomes (**Supplement 1**). Therefore, as time post-injury increased, there were modest increases in minutes of daily MPA and VPA, but not sedentary time or LPA.

## DISCUSSION

This is the first study to characterize habitual physical activity and sedentary time using accelerometry in children with concussion in comparison to healthy controls. We report that children with concussion are significantly more sedentary and less active (with respect to LPA, MPA, and VPA) than matched healthy controls; this is observed throughout the day, from 8:00AM to 9:00PM. Sex-specific analyses showed that girls with concussion are less active (across all intensities studied) and more sedentary than boys with concussion as well as healthy girls. Increased days post-injury significantly predicted higher levels of MPA and VPA–but not sedentary time or LPA–in children with concussion, which may be expected as a natural part of concussion recovery.

### Prolonged inactivity in concussion

We identified a mean difference of 38.3 minutes/day (95% CI, 11.2, 65.4) of increased sedentary time in children with concussion compared to their healthy peers, which amounts to nearly 4.5 hours a week. This difference is even greater when comparing girls with concussion to healthy girls, where the mean difference in sedentary time is 51.1 minutes/day, or approximately 6 hours over a week. Concussion guidelines now suggest that prolonged rest should be avoided after the first 24-48 hours of injury^1^, and a recent randomized trial^25^, large-scale cohort study^24^, and systematic review^19^ support this notion. However, what constitutes prolonged rest (or sedentariness) is not yet well-defined, as indicated in the most recent sport concussion guidelines^1^. Our data do not establish a definition of prolonged rest post-concussion, but they do quantify the magnitude of the physical activity observed within the first month of injury. Instead of (or in addition to) prescribing a designed 20-minute period of aerobic activity as is current best practice^14-16^, advising patients to interrupt their sitting may be advantageous, especially given the known benefits of such actions on multiple health outcomes in healthy and other clinical populations^39-41^. The optimal amount of sedentary time and physical activity following concussion, and whether a single bout of activity has greater benefits than interrupted sitting, requires further study. It is also important to note that sedentary time is not the inverse of physical activity, with each having different physiological effects^42^. The contribution of each to concussion outcome needs further study.

### Engaging key stakeholders in the exercise discussion

With the accumulation of data on the benefits of exercise in concussion, the traditional “rest is best” approach is being overturned, with an “exercise is medicine” mindset becoming more commonplace. However, a knowledge translation gap still exists, wherein primary care providers have not yet adopted this new approach to concussion management; more than 80% of concussion patients are still advised to rest for more than 2-days, despite contrary evidence from recent guidelines and reviews^1,17,20^. A recent study aimed at providing primary care providers with guidelines for de-implementing prolonged rest found that the intervention improved knowledge about avoiding prolonged rest post-concussion, and increased clinician adherence to guideline recommendations from 25% to nearly 90%^43^. Further, per research on military service members with concussion, primary care providers relaying information to patients about the consequences of prolonged rest led to patients more promptly resuming physical activity and self-reporting lower levels of symptoms sooner^44^. Future research should not only continue to build on the evidence, but also ensure that key findings are translated to relevant knowledge users.

### Towards F.I.T.T. informed exercise interventions in concussion

The bulk of exercise research in concussion has been on sub-maximal aerobic exercise^45^. Studying the impact of low-to-moderate intensity aerobic activity was motivated by the desire to engage patients in a safe, symptom-limited amount of activity that had a high likelihood of being well-tolerated. The optimal exercise frequency, intensity, time, and type (F.I.T.T.) of exercise in concussion has not been directly studied, though such study is necessary to maximize clinical benefit and increase exercise adherence in clinical populations^46^. Recently, however, research has started to provide insight into F.I.T.T principles for exercise prescription in concussion. Accelerometer studies show that engaging in MVPA following concussion can lead to longer recovery times^32^, which supports the continued use of low-to-moderate intensity graded sub-maximal aerobic exercise programs at this time^15^. Further, in adolescents, participating in low-intensity aerobic exercise for less than 100 minutes/week was associated with greater symptom burden at one-month post-injury, while exercising more than 160 minutes/week resulted in symptom resolution when assessed at the same timepoint^47^. These data, in addition to the current findings that quantify the physical activity debt observed in pediatric concussion, are helpful for building towards an understanding of the optimal frequency and time of exercise interventions in concussion. Our data (**Supplement 1**) also show that the effect sizes for the difference in sedentary time between children with concussion and healthy controls is greatest from 8:00 AM to 1:00 PM, and then again from 8:00 PM to 9:00 PM. Given that sedentariness is observed throughout the day, but particularly so during school-going hours, there may be opportunity to incorporate physical activity programs as part of return-to-learn programs.

### Accelerometery in concussion-exercise research

Despite the recognized importance of physical activity on concussion symptoms, accelerometer research in pediatric concussion has been limited. To date, studies have used accelerometry to assess the relationship between physical activity with subsequent outcome in non-controlled cohort studies. More specifically, one study showed that the number of accelerometer-measured steps from days 1-3, 4-5, and 6-7 post-injury were significantly correlated with symptom scores at each of these intervals, and the increased activity (i.e., number of steps) from days 1-3 post-injury predicted lower activity on days 4-5 post-injury^33^. A second study by this group reported that there was no association between physical *and* cognitive activity and time to concussion recovery^34^. Another group studied male youth hockey players wearing accelerometers in the early stages of injury and found that those in the “high” activity group (based on a median split, performing more than 148.5 minutes of MVPA/day) took significantly longer to recover than those in the “low” activity group^32^. Our study, in comparing accelerometer-measured sedentariness and activity between cohorts of concussed and healthy children, shows that there are considerable differences in sedentary time and physical activity within the first-month of injury. Future research should continue to use accelerometers over self-recall questionnaires to build on these insights. Importantly, self-report measures of physical activity have been shown to under-estimate sedentary time and over-estimate activity in comparison to device-measured physical activity behaviours in both children^48,49^ and adults^49,50^. Moreover, adopting accelerometry broadly in concussion research creates opportunities for data sharing and big data analysis, recognized as an important aspect in the next wave of neuroscience research^51,52^.

## LIMITATIONS AND FUTURE DIRECTIONS

Our study is limited by the absence of symptom data that preclude exploration of the association between accelerometer-measured physical activity and clinical outcome. Participants in our study also wore the accelerometer over their right hip; whether there are differences in measured activity based on location of wear remains unknown. Further, for our sedentary bout analysis, we used a 6-second drop-time as this length of time was estimated to not be physiologically-relevant yet akin to slight movement such as repositioning or reaching for a device. Drop time criterion can be better established by future research.

Additional research is required to understand the threshold of total volume or number of bouts of sedentary time beyond which concussion recovery is compromised. Alternate accelerometer-based physical activity metrics, including indices of movement variability, can also provide more insight into how activity patterns differ in children with concussion in comparison to their healthy peers. Cohort studies and longitudinal research examining how acute clinical features of the injury associate with subsequent physical activity are needed for identifying children at high risk of sedentariness post-concussion.

## CONCLUSIONS

This is the first study to profile accelerometer-measured physical activity and sedentariness following pediatric concussion, finding that in the first-month of injury children with concussion are more sedentary (MD, 38.3 minutes/day) and less active (across all intensities studied) than their healthy peers. We also report that as time post-injury increases, levels of MPA and VPA increase in children with concussion, which may be consistent with natural recovery. Future exercise-concussion research should examine the impact of interventions that reduce sedentary time, and in general engage key stakeholders (including primary care providers) to improve knowledge translation and the adoption of exercise in clinical practice.

## Data Availability

Data are available upon reasonable request.

## STATEMENTS

### Contributorship

BW Timmons, MD Noseworthy, and C DeMatteo were involved in conceptualizing, planning, and securing funding for the parent study. C DeMatteo maintained study oversight over the parent study. J Obeid provided oversight over accelerometer data collection and analyses. B Sharma defined the research question for this secondary data analysis, cleaned and analyzed all accelerometer data, and prepared the first draft of the manuscript.

## Acknowledgements

We would like to thank all study participants for their time and commitment to this research.

## Funding, grant, and award info

This research was supported by the Canadian Institutes of Health Research (#31257), as well as Doctoral support to B Sharma from the Canadian Institutes of Health Research (CIHR-CGS-D, #157864). BW Timmons is the Canada Research Chair in Child Health & Exercise Medicine.

## Ethical approval information

The study was approved by the Hamilton Integrated Research Ethics Board (www.hireb.ca) # 14-376.

## Data sharing agreement

Data are available upon request.

## Patient involvement

Given that this study is a secondary data analysis, patient and public involvement was not part of this analysis. For the parent study, however, patients and the public: were involved with research at the time of recruitment; did not inform the research question or outcome measures; were not involved in study design; were actively recruited as community-based research participants; were informed of the time requirements of the study; and were active in the knowledge translation plan.

